# A systematic review and meta-analysis of the incidence of breast cancer-related lymphoedema due to treatment combinations

**DOI:** 10.1101/2025.06.11.25329404

**Authors:** Jennifer Frattolin, Iva Grujic, Arushri Swarup, Alain Vella, Ilias Kalamaras, Bingxuan Wang, Daniel J. Watson, Anastasios Drosou, Paul Thiruchelvam, James E. Moore

**Affiliations:** University of British Columbia, School of Engineering, Faculty of Applied Science, Kelowna, Canada; Department of Bioengineering, Imperial College London, London, United Kingdom; Information Technology Institute (ITI), Centre for Research and Technology Hellas (CERTH), Thermi, Thessaloniki, Greece; Charing Cross Hospital Campus, Imperial College Healthcare NHS Trust, London, United Kingdom

## Abstract

**BACKGROUND:** Breast cancer related-lymphoedema (BCRL) is a chronic, debilitating disease for which there is no cure. A meta-analysis was conducted to estimate the association between different treatment combinations on the incidence of BCRL.

**METHODS:** The review was conducted according to PRISMA guidelines with four databases searched for studies published from 2000-2020, including OVID Medline, OVID Embase, Cochrane Library for Registered Controlled Trials, and Cumulative Index to Nursing and Allied Health, yielding 2640 studies. A random effects model was used to determine BCRL incidence rates stratified by treatment types of the qualifying studies.

**RESULTS:** The pooled incidence rate was 23% (95% CI 20.8 – 25.4) for patients who received axillary lymph node dissection (ALND) and 5.6% (95% CI 4.5 – 6.8) for patients who underwent sentinel lymph node biopsy (SLNB). A higher level of intervention to the axilla was identified as the key factor associated with significantly increased BCRL incidence, including ALND (p<0.001), the number of lymph nodes removed (p<0.001), and axillary radiotherapy (p<0.001). Higher patient BMI was also identified to increase BCRL incidence. Combinations of other treatments, in conjunction with ALND or SLNB, did not lead to statistically significant differences in incidence. Furthermore, different diagnostic criteria resulted in substantial variation in BCRL incidence rates.

**CONCLUSION:** Axilla intervention was associated with increased BCRL incidence including ALND, number of lymph nodes removed, and radiotherapy. The inclusion of additional surgical or non-surgical treatments did not. This analysis re-emphasises the clear need for standardised reporting of patient treatments as well as universally applied diagnostic protocols.

## 1. Introduction

Breast cancer-related lymphoedema (BCRL) is a debilitating condition that can develop following treatment for breast cancer. It results in arm swelling, impaired function, chronic infections, and impedes return to a functional lifestyle (1). BCRL is also a constant physical reminder of cancer and makes patients prone to anxiety and depression (2, 3). A reported 2.26 million new cases of female breast cancer were diagnosed worldwide in 2020 (4), with incidence rates rapidly increasing in developing countries (5). As more targeted breast cancer treatments increase long-term patient survival, with a 10-year survival rate of 80% (6, 7), understanding the influence of the various factors that increase incidence of BCRL is important for both patients and healthcare systems.

Cancer treatments that influence BCRL incidence include the nature of surgical and non-surgical procedures, spanning axillary lymph node dissection (ALND), sentinel lymph node biopsy (SLNB), number of lymph nodes (LNs) removed, mastectomy, and lumpectomy (8–11). Additional treatments such as radiotherapy (RT), chemotherapy (CT), and hormone therapy (HT) may increase incidence (12, 13). Clinical patterns in the application of these treatments have evolved, with a trend towards de-escalation of surgical invasiveness (14), and there have been trials to investigate the effects on patient survival (15). Given that consideration of BCRL played a role in the movement away from ALND and towards SLNB in particular, one might expect that BCRL incidence rates should be declining.

Other factors that influence BCRL incidence and may therefore make procedural effects more difficult to detect include patient characteristics such as genetic predisposition, racial origin, sedentary lifestyle, and BMI(16–23). Further impeding detection is the fact that BCRL diagnosis depends on the diagnostic technique and criteria applied (24, 25). Lastly, the time course associated with BCRL onset varies from months to years, so a minimum follow-up time of 24 months is essential but not always followed (10, 26, 27).

The most recent meta-analysis of BCRL incidence rates was published by DiSipio et al. in 2014 (28). They surveyed 398 potentially relevant studies, 72 of which were chosen for further analysis. In this meta-analysis, incidence rates were stratified by patients who received SLNB or ALND only, with rates of 5.6% and 19.9%, respectively. To address this gap in understanding of how treatment combinations affect BCRL incidence rates, we performed a new systematic review and meta-analysis of BCRL incidence based on treatment combinations, which has not been previously undertaken. The aim of this work was to provide insight into the effects of treatment combinations, patient characteristics, and diagnostic technique on BCRL incidence. Ultimately, we seek to provide breast cancer patients, clinicians, and researchers with an updated knowledge base of BCRL incidence to increase awareness of this debilitating, incurable disease.

## 2. Methods

### 2.1. Search strategy and selection criteria

A systematic review was conducted to identify all studies that report BCRL incidence as a result of the treatment(s) administered to patients (Fig. 1). The review was conducted in collaboration with an experienced librarian, according to PRISMA guidelines, and registered on PROSPERO (ID No. CRD42022309978). Four databases were searched: OVID Medline, The Cochrane Library for Registered Controlled Trials, Cumulative Index to Nursing and Allied Health, and OVID Embase, using the following search terms: breast cancer (“breast cancer”, “breast neoplasms”, or “breast carcinoma”), lymphoedema (LE) (“lymphoedema”, “lymphedema”, “breast cancer lymphedema”, “arm swelling”, or “arm edema”), and treatment (“sentinel lymph node biopsy”, “lymph node excision”, “axillary dissection”, “axillary surg”, “axillary node dissection”, “drug therapy”, “chemotherapy, “radiotherapy”, “radiation”, “hormone therapy”, “mastectomy”, “lumpectomy”, “breast conserving surgery”, or “risk factors”). Search results were restricted to studies published between January 1, 2000, and December 31, 2020. This cutoff date was chosen because publications after this time frame may have contained data influenced by the COVID-19 pandemic. A total of 2640 studies were identified for screening for which the systematic review management software, Covidence (Melbourne, Australia), was used.

**Fig. 1.**
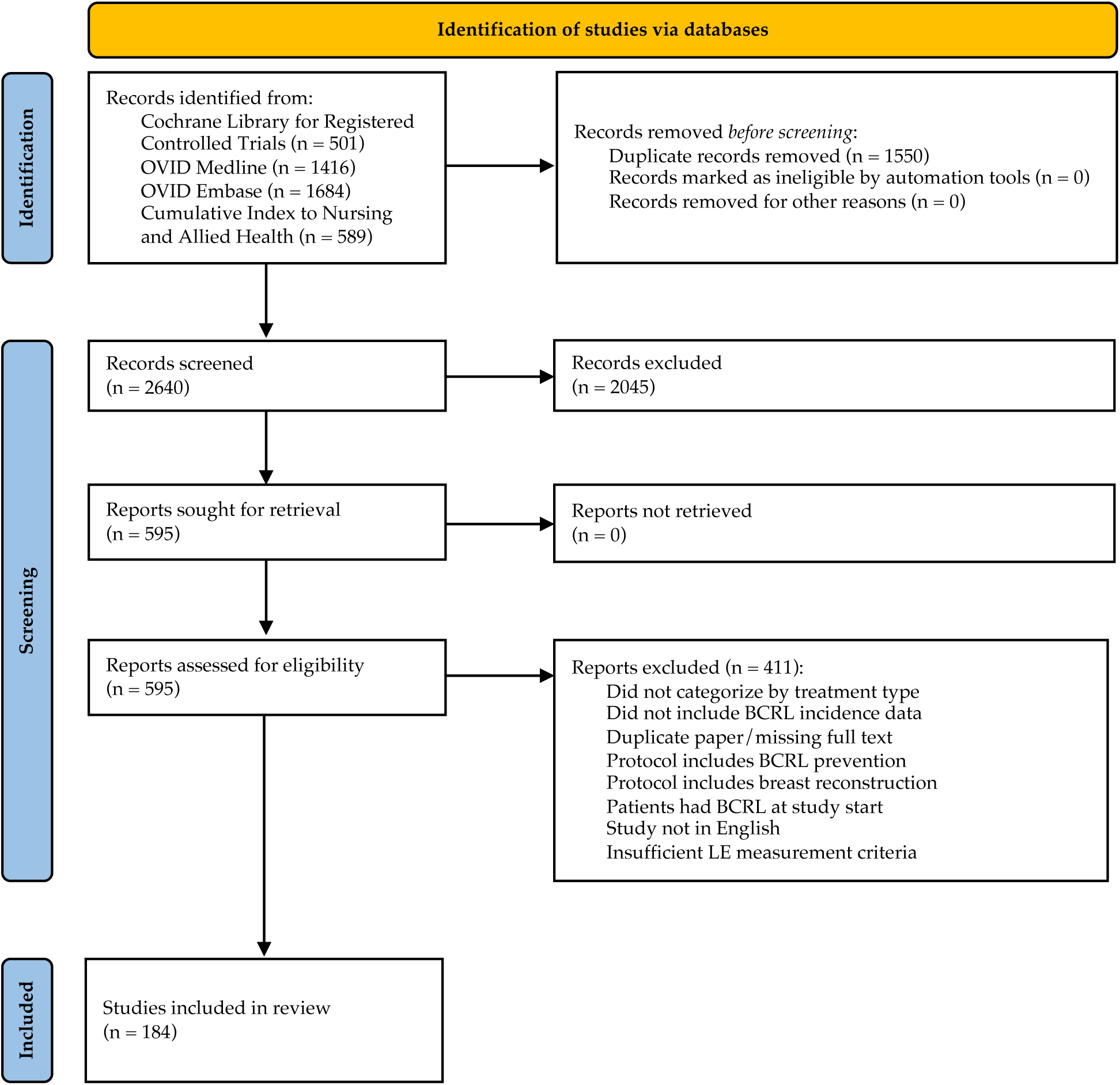
PRISM flowchart of the systematic review process. A total of 184 articles were selected for inclusion and data extraction from an initial screening of 2640 studies.

The categories for eligibility criteria for meta-analysis inclusion were study type and language, patient characteristics, design/outcome, and diagnostic method. For the study type and language, only published research articles were included, while review papers, meta-analyses, editorial or comment papers, case reports, and case series were excluded. Research articles not published in English were excluded. Studies including female patients with either unilateral or bilateral breast cancer, without a previous diagnosis of BCRL, were included. Studies that investigated LE prevention techniques on patient BCRL incidence were excluded to avoid biasing the dataset. In addition, patients who underwent breast reconstruction at the time of surgery were excluded; a meta-analysis by Siotos *et al.* in 2018 indicated that breast reconstruction at the time of breast cancer surgery significantly reduced BCRL incidence (29). Eligible studies were required to stratify incidence with respect to at least one of the following treatments: ALND, SLNB, mastectomy, lumpectomy, RT, CT, or HT. In addition, BCRL incidence must be reported as the number of patients with BCRL within the study or as a rate of incidence. Lastly, the criteria and/or method for BCRL assessment and diagnosis must be specified for the study to be included. All objective and subjective methods to diagnose LE were accepted for this meta-analysis, including bioimpedance spectroscopy, arm circumference and volume measurements, lymphoscintigraphy, clinician evaluation, and patient self-assessment.

### 2.2. Data extraction

Initial screening of the title and abstract was conducted in Covidence based on the eligibility criteria described to select studies for full-text review. Each article was assessed by two reviewers and conflicts were resolved by a third reviewer (A.S., A.V., I.G., and J.F.); this process excluded 2045 studies from the analysis. Full text review of the remaining 595 studies followed a similar review procedure of which 184 articles were included for data extraction (Fig. 1).

The following data were extracted: BCRL incidence per patient group, the treatments that were and were not administered (ALND, SLNB, mastectomy, lumpectomy, RT, CT, and HT), study year, location, and type, as well as the method of BCRL assessment/diagnosis. In this analysis, the following surgical treatments were considered the same and referred to as lumpectomy: breast conserving surgery, lumpectomy, and wide local excision. When extracting incidence data per patient group, a binary classification was used to identify which of the 7 treatments were administered, with ‘1’ indicating if a treatment was received and ‘0’ if a treatment was not. If it was unknown whether a treatment was given, it was identified with ‘N/A’. The following data were also extracted from each study when available: the number of LNs removed, the type of chemotherapy received, the location of radiotherapy, and patient BMI. The data were extracted by J.F., A.S., A.V., and B.W. and verified by J.F., I.G., and B.W. for data accuracy and duplication.

### 2.3. Statistical analysis

The extracted incidence data were categorised by treatment type. ALND and SLNB were directly compared in combination with surgical and non-surgical treatments. We defined a surgical (S) treatment as at least one instance of mastectomy or lumpectomy. Similarly, at least one treatment of CT, RT, or HT was required to qualify as a non-surgical (NS) treatment. A total of 8 treatment categories were possible: ALND, ALND + S, ALND + NS, ALND + S + NS, SLNB, SLNB + S, SLNB + NS, and SLNB + S + NS. For clarity, ALND + S refers to all BCRL incidence rates associated with ALND and surgical treatments but excludes ALND associated studies that did not report surgical treatments. The categorisation process followed for all extracted data is illustrated in a flowchart (Fig. 2). This flowchart was also designed to prevent potential patient overlap within groups (Fig. 2), which was associated with incomplete reporting of patient treatments administered.

**Fig. 2.**
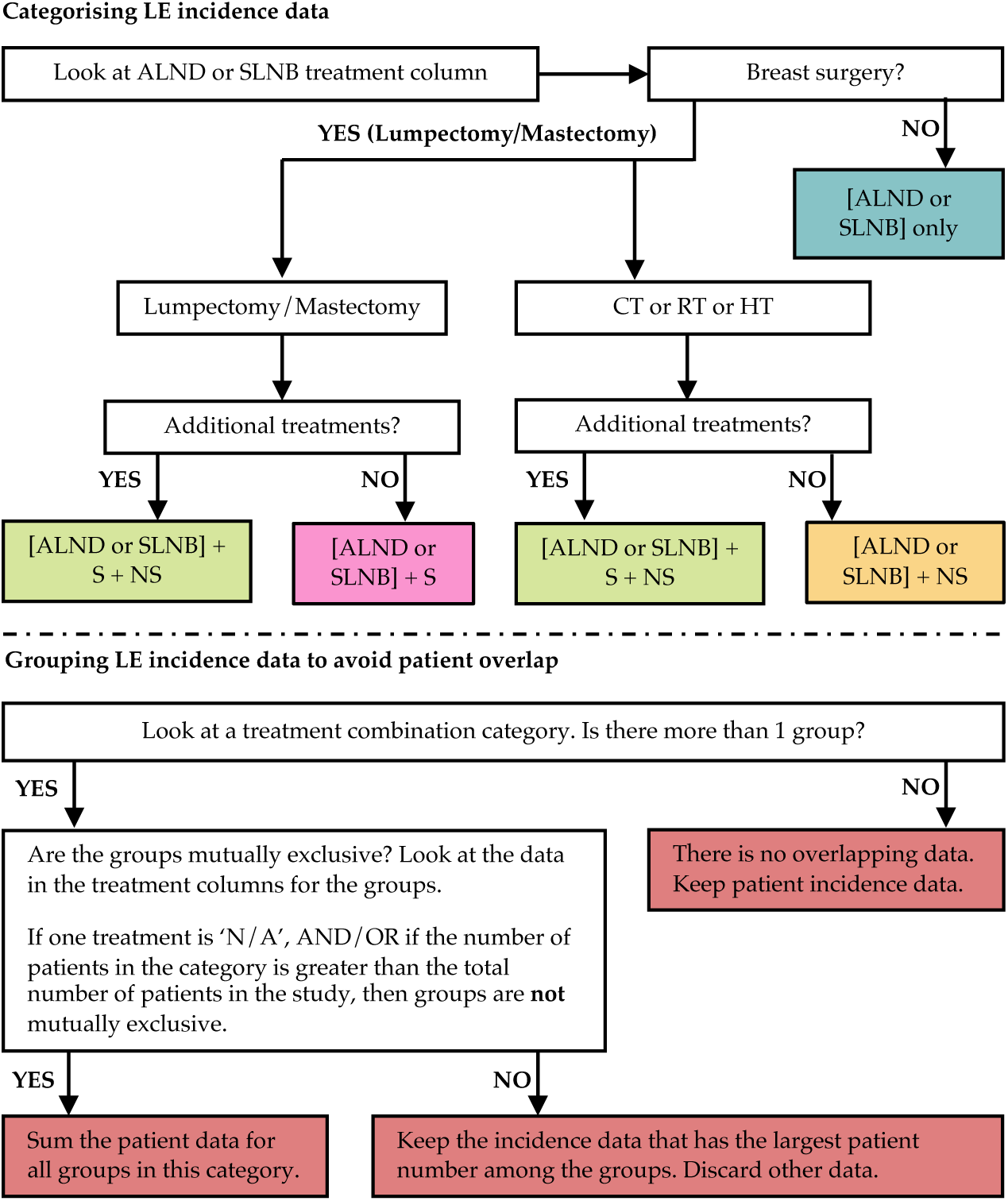
Decision flow chart developed to consistently group incidence data into treatment categories as well as to prevent potential patient overlap within a treatment category. Shaded boxes within the flowchart refer to a finalised result. S: surgical treatments (mastectomy, lumpectomy; NS: non-surgical treatments (RT, CT, HT).

A random effects model using the ‘metafor’ package within R (The R Foundation, New Zealand) was implemented for the meta-analysis. This model was selected to help account for potential variation between groups and unobserved heterogeneity within the dataset. Incidence rates and 95% confidence intervals for each treatment category were calculated, with the effect size considered to be the mean incidence rate. Publication bias was assessed using the Egger’s test. Comparisons between groups were conducted using a two-sample weighted t-test with a Bonferroni correction to account for multiple comparisons. A p-value of less than 0.05 was considered to be significant. Additional sub-analyses were conducted in a similar manner to evaluate how specific patient, treatment, and study characteristics influenced BCRL incidence rates. These include the number of LNs surgically removed, RT location, CT type (adjuvant vs. neoadjuvant), patient BMI, and methods to diagnose LE. Kernel density estimates were used to investigate how SLNB and ALND incidence rates have changed during the time period for qualifying periods. Certainty of evidence was assessed using the GRADE approach was implemented according to Cochrane Reviews (30).

## 3. Results

### 3.1. Breast cancer treatment combinations

A total of 184 studies were identified for inclusion (Supplementary Material), including prospective cohort (n=69), retrospective cohort (n=66), cross-sectional (n=18), randomised clinical trials (n=21), and comparative (n=7) studies, as well as one each of longitudinal, populational, and preclinical studies. Studies were synthesized by treatment combination including ALND only, ALND + S, ALND + S + NS, SLNB only, SLNB + S, and SLNB + S + NS and mean incidence rate determined (Fig. 2). The result of each individual study is provided in the Supplementary Material. Grouping the results by treatment type yielded an incidence rate of 5.6% (95% CI 4.5-6.8) for all SLNB groups and 23% (95% CI 20.8-25.4) for ALND groups. A two-sample weighted t-test found the pooled SLNB and ALND incidence rates to be significantly different (p<0.0001, high certainty of evidence). Paired treatment combinations, such as SLNB + S and ALND + S, were significantly different (p<0.01) from each other for all combinations in the analysis (Fig. 3). This identifies the critical impact of ALND on BCRL incidence, regardless of what other treatments are performed. The addition of further treatments, including surgical and non-surgical, did not significantly affect BCRL incidence within the SLNB and ALND categories (moderate certainty of evidence, downgraded for potential publication bias). Furthermore, stratification of the data based on study type, specifically prospective cohort studies only, did not yield significantly different results.

**Fig. 3.**
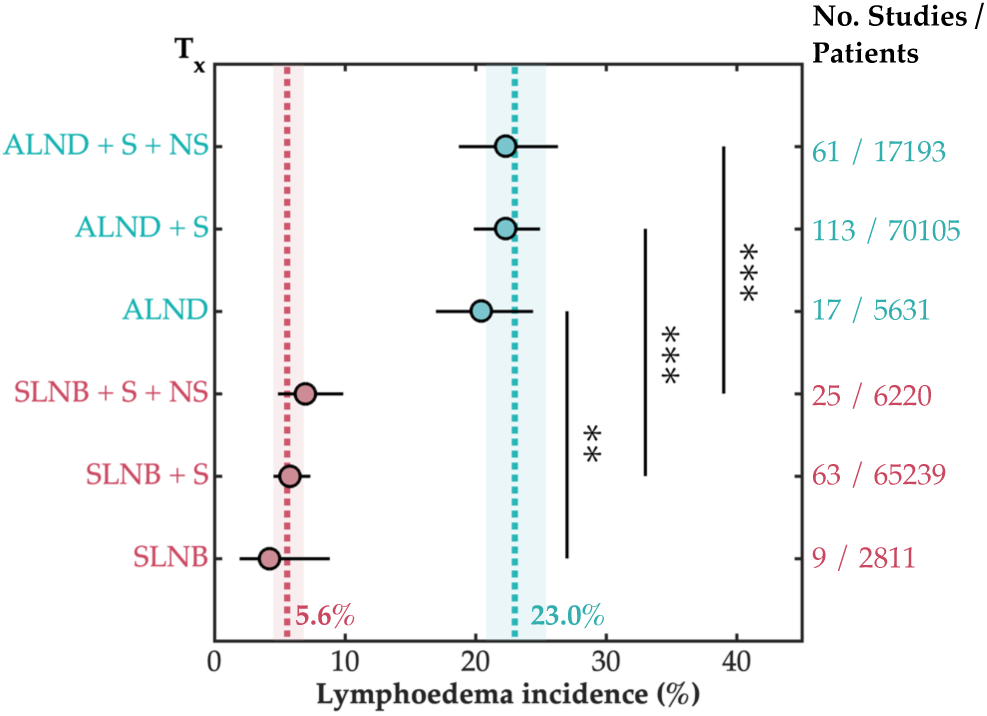
Results of the meta-analysis comparing SLNB and ALND plus treatment types, including surgical (S) and non-surgical (NS) treatment options (T_x_). The mean incidence is reported for each category with the 95% confidence interval. The number of studies and patients included in each treatment category are indicated on the right. The overall incidence rate for SLNB, indicated by the dashed pink line, was significantly lower than that of ALND-based treatments, indicated by the dashed blue line (p<0.001). Limited studies existed for the potential combination of SLNB + NS and ALND + NS and were thus excluded. (NB: p<0.01 is denoted with (**), and p<0.001 is denoted with (***)).

Publication bias was assessed using an Egger’s test for each treatment combination category. The following categories indicated potential bias upon failing the Egger’s test with p<0.05, including ALND + S, ALND + S + NS, and SLNB + S + NS. All other categories were not found to have bias. To investigate this further, a sensitivity analysis was conducted (Section 3.1.1).

#### 3.1.1. Sensitivity analysis of breast cancer treatment combinations

In the process of compiling incidence data based on breast cancer treatment combinations, it was not assumed that a patient cohort received any treatments other than what was explicitly reported within the study. As a result of this assumption, multiple surgical or non-surgical treatment types were grouped together. This results in a decrease in granularity of the dataset, and it is hypothesised that this is the source of the potential bias. To assess if this significantly modified the results of the meta-analysis, the analysis was conducted only with studies that were classified as ‘complete’; this was equal to 30 studies that did not have any missing patient treatment data.

Stratifying by treatment combinations yielded a BCRL incidence rate of 26.5% (95% CI 19.5-32.9, n=21) and 8.0% (95% CI 5.6-11.5, n=21) for the ALND + S + NS and SLNB + S + NS groups, respectively, of this dataset. ALND/SLNB + S only had one study each and were excluded; no studies remained of the ALND/SLNB only category. Two-sample t-tests were used to compare incidence rates between the ‘complete’ datasets and those presented in Fig. 3. It was determined that the SLNB + S+ NS and ALND + S + NS categories were not significantly different from its corresponding category of Fig. 3, p=0.4933 and p=0.5745, respectively. It was concluded then that this source of bias resulting from the decreased granularity of the grouping categories did not significantly alter the results.

### 3.2. Change in SLNB- and ALND-stratified LE incidence by publication year

The relative change in ALND and SLNB-stratified incidence was assessed over the 20-year period of this meta-analysis. Kernel density estimations were used to evaluate probable LE incidence distribution over time (Fig. 4). There is an increase in the reported LE incidence associated with ALND treatment. Incidence associated with SLNB treatment has remained relatively constant for the past ten years.

**Fig. 4.**
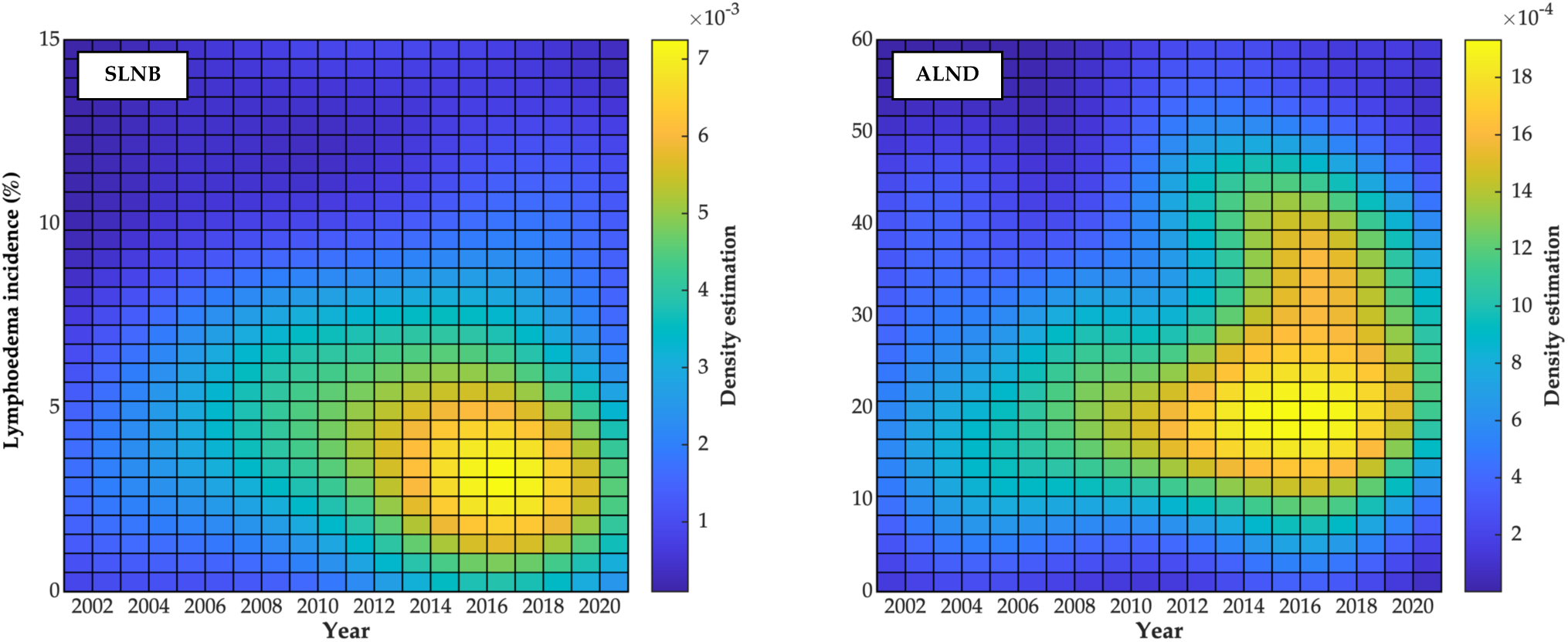
The kernel density estimations of BCRL incidence were determined for SLNB and ALND from 2001 to 2021. An increase in incidence is observed for ALND after 2015, while SLNB-related BCRL incidence remained relatively constant.

### 3.3. Number of lymph nodes removed

BCRL incidence was further stratified by the number of LNs (N_LNs_) removed based on a subset of 32 studies that included these data (Fig. 5a). Publication bias for this sub-analysis was evaluated using an Egger’s test with no significant bias found (p>0.05 for all categories). Increased incidence was observed when more than ten LN were removed, 24.5% (95% CI 21.3-28.1), compared to an incidence of 15.2% 24.5% (95% CI 21.3-28.1) when less than ten LNs were removed. A two-sample t-test comparing incidence rates of N_LNs_ < 10 and N_LNs_ > 10 was statistically significant (p<0.0001, high certainty of evidence). Incidence did not significantly increase further when > 10 LNs were removed.

**Fig. 5.**
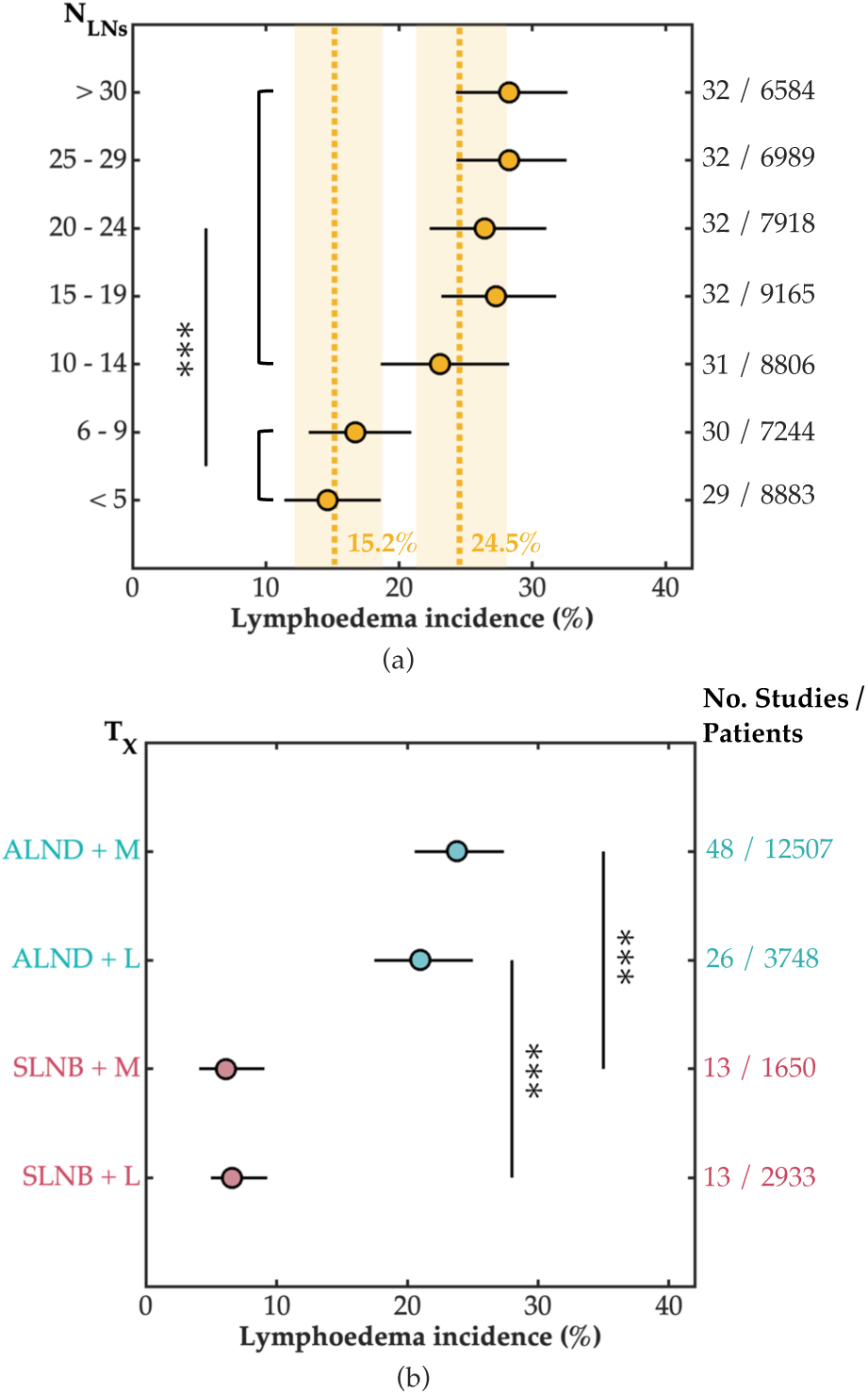
(a) BCRL incidence as a result of the number of LNs (N_LNs_) removed during axillary surgery, with mean incidence reported and its corresponding 95% confidence interval. A significant jump in incidence was observed in studies where ten LNs or more LNs were removed (p<0.0001), after which incidence did not increase significantly. (b) ALND/SLNB stratified incidence for mastectomy (M) and lumpectomy (L) patients, with mean incidence reported and its corresponding 95% confidence interval. No significant difference was observed in incidence whether a patient received mastectomy or lumpectomy.

### 3.4. Breast surgery

Analysis of individual surgical treatments showed that BCRL incidence was significantly higher for ALND patients than SLNB patients (p<0.0001), regardless of whether they received a mastectomy or lumpectomy (Fig. 5b). There was an incidence of 23.8% (95% CI 20.5-27.4) vs. 21% (95% CI 17.5-25.0), for ALND patients who received mastectomy and lumpectomy, respectively, in contrast to 6.1% (95% CI 4.1-9.1) and 6.6% (95% CI 4.8-9.1) for SLNB patients. Incidence was not significantly increased if a patient received mastectomy over lumpectomy for either ALND or SLNB patients (moderate certainty of evidence, downgraded for potential publication bias). The results of the Egger’s test for this sub-analysis revealed potential for bias in the SLNB + Lumpectomy category (p=0.0047); the results from all other categories did not (p > 0.05).

### 3.5. Non-surgical treatments

Other treatment subtypes, such as RT location, had a significant effect on BCRL incidence (Table 1), with axillary RT resulting in the highest incidence of 31.1%. BCRL incidence of axillary RT were significantly higher in comparison to chest/breast RT (p<0.0001, moderate certainty of evidence downgraded for potential publication bias). The time course of CT, either adjuvant or neoadjuvant, did not have a statistically significant effect on incidence with p=0.095 (high certainty of evidence). Publication bias for these sub-analyses was assessed using Egger’s tests, where no publication bias was found for CT dataset. For RT location dataset, the results of the Egger’s test revealed potentially bias in the chest/breast subcategory (p=0.0082), while the axillary RT did not (p=0.90).

**Table 1.** BCRL incidence rates resulting from different treatment options including radiotherapy location and chemotherapy time course.

| Treatment |  | Lymphoedema incidence,<br>%<br>(95% CI) | Number of studies /<br>patients |
| --- | --- | --- | --- |
| Radiotherapy | Axillary | 31.1 (23.8 – 39.4) | 19 / 1110 |
|  | Chest/breast | 12.5 (9.53 – 16.2) | 36 / 17596 |
| Chemotherapy | Neoadjuvant | 25.9 (19.2 – 34.1) | 22 / 5018 |
|  | Adjuvant | 17.8 (12.6 – 24.7) | 18 / 4830 |

### 3.6. Patient BMI

Combining the results of studies containing data on patient BMI confirmed that increased BMI led to a greater incidence of BCRL (Table 2, high certainty of evidence). Publication bias for the sub-analysis was assessed using an Egger’s test with no significant bias found (p>0.05 for all categories). A statistically significant increase in LE incidence was observed between BMI of less than 24.9 and BMI of greater than 30 (p=0.0012). However, the initial increase in LE incidence was observed at a BMI of 25 – 29.9, though this was not statistically different (p=0.055). A BMI > 30 was not found to be statistically different from that of a BMI between 25 – 29.9 (p=0.72).

**Table 2.** BCRL incidence increased with higher BMI. A jump in BCRL incidence was observed with an overweight BMI in comparison to those with a BMI < 24.9, but minimal difference was observed once patient BMI exceeded 25.

| BMI | Lymphoedema incidence, %<br>(95% CI) | Number of studies / patients |
| --- | --- | --- |
| < 24.9 | 16.7 (13.7 - 20.1) | 34 / 14146 |
| 25 - 29.9 | 23.8 (19.2 - 29.1) | 33 / 10476 |
| > 30 | 28.3 (23.2 - 34.1) | 36 / 8368 |

### 3.7. Methods to diagnose lymphoedema

A broad range of BCRL diagnostic timings and methods were reported, with the time of LE diagnosis defined by the number of years since treatment or cancer diagnosis. This varied greatly from 2 months to 20 years, with a right skewed distribution (Fig. 6a). A median of 2.9 years was identified; continued observation of at least 24-36 months has been recommended (26, 27, 31).

**Fig. 6.**
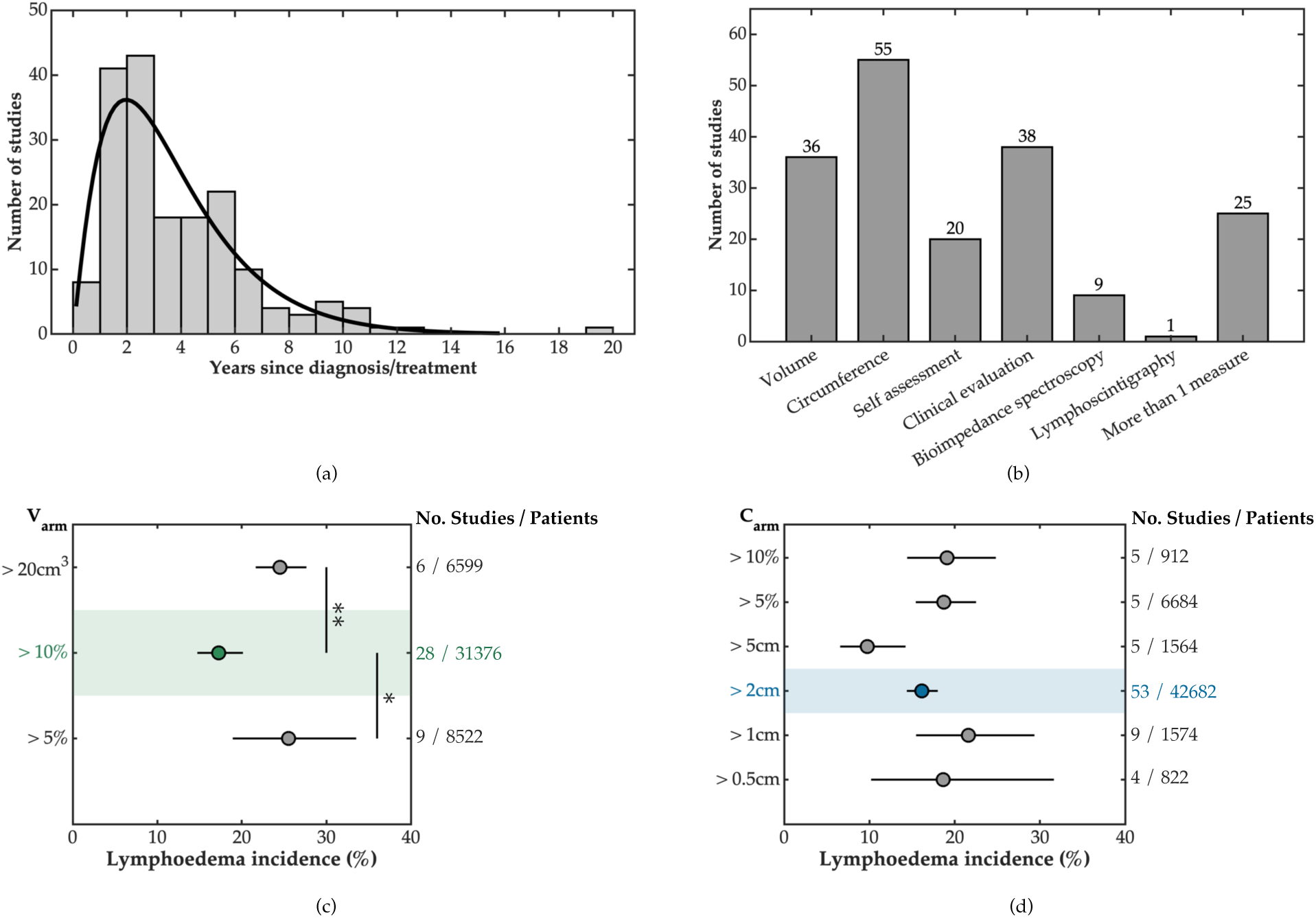
(a) Mean/median time since breast cancer diagnosis or treatment for all studies in the meta-analysis with a gamma distribution fitted. Numbers do not add up to 184 (100% of included studies) as some studies had multiple follow-ups or only provided a range of follow-up periods and were thus excluded. (b) Six different methods to diagnose BCRL were identified, with arm circumference being the most prevalent. Volume-based methods included water displacement measurements, the frustrum method, and perometry. More than one measure of diagnosis refers to one subjective measure (patient self-assessment or clinical evaluation) and one objective measurement (volume, circumference, or bioimpedance), or two objective measures. BCRL incidence rates comparing different clinical diagnostic thresholds for both (c) arm volume and (d) arm circumference, with mean incidence reported and its corresponding 95% confidence interval. The shaded bands, in green and blue, represent the literature consensus as the diagnostic criterion for both circumference and volume-based measurements, respectively. (NB: p<0.05 is denoted with (*), and p<0.01 is denoted with (**)).

Six different methods were used to diagnose BCRL (Fig. 6b), with circumference measurements being the most prevalent method followed by patient self-assessment and clinical evaluation. Clinical evaluation was defined as a diagnosis of BCRL following a clinician, nurse, or physiotherapist assessment, or a retrospective analysis of patient records. Within both circumference and volume-based measurements, a range of diagnostic criteria was employed. Significant differences were observed between volume-based criteria (Fig. 6c, high certainty of evidence). The most commonly used metric, a 10% increase in volume, yielded an incidence of 17.3% (95% CI 14.7-20.1). This was statistically lower than incidence using a volume increase of 20cm^3^, 24.5% (95% CI 21.6-27.7), and a 5% increase in arm volume, 25.5% (95% CI 18.9-33.5). The results of the Egger’s test revealed no publication bias (p > 0.05).

In contrast, there was no significant difference in incidence between the most frequently used circumference diagnostic metric of 2cm increase and any other circumference-based measurement (moderate certainty of evidence, downgraded due to potential publication bias), with wide variations in mean values (Fig. 6d). The Egger’s test of the circumference-based measurements revealed potential publication bias for the 2cm circumference diagnostic metric (p<0.0001).

## 4. Discussion

This meta-analysis presents a current trend on BCRL incidence rates and the effect of different treatment combinations on its incidence. More invasive interventions to the axilla increased BCRL incidence, including the use of ALND, the number of the LNs removed, and axillary radiotherapy. Irrespective of treatment history, increased patient BMI correlated with a higher incidence of BCRL. BCRL incidence was over four times greater for patients who received ALND versus only SLNB. Kernel density analysis demonstrated that LE incidence associated with ALND treatment has risen over the past ten years. This increase is likely attributed to changes in clinical practice; whilst fewer ALND procedures are undertaken overall (32), those patients tend to have more advanced disease and are more likely to receive complex therapy, including removing a higher number of lymph nodes.

Today, breast-conserving surgery and axillary surgery de-escalation are the gold standard for breast cancer treatment in developed countries. According to the European Society of Medical Oncology (ESMO), 60-80% of new breast cancer patients in Western Europe are eligible for a breast-conserving treatment approach (33). This is in contrast to a century ago, where the Halsted’s procedure was routine practice, which removed the breast, axillary LNs, and chest muscle resulting in significant patient morbidity and disability (34). Axillary surgery de-escalation, marked by the development of SLNB in the 1990s (35), followed breast conserving surgery as a further means to reduce patient morbidity. SLNB is now the standard of care for axillary staging in early, clinically node-negative breast cancer, in part due to the ACOSOG Z0011 (Alliance) randomized clinical trial, which found that routine use of ALND did not improve patient outcomes in low burden disease (36). Further to this, in 2019, additional axillary intervention for micro-metastases or 1-2 sentinel node macro-metastases was no longer recommended across the UK, Europe, United States, and Canada (33, 37, 38). According to the National Cancer Database in the United States, 77% of the 1.13 million patients diagnosed with Stages I-III breast cancer from 2004-2014 underwent non-ALND management (32).

The change in practice towards axillary de-escalation does not represent current clinical practice in every healthcare system, however. In India and China, SLNB may not be offered in all medical centres or by all surgeons due to training deficiencies, limited access to radioisotopes, and financial restrictions (39). A recent study by Zhang *et al.* revealed that only 43.5% of early-stage breast cancer patients received SLNB in China, with surgeons preferring ALND in single node-positive patients (40). This is in stark contrast to the SEER database review by Gou *et al.*, where 81% of patients in the United States received SLNB in 2016 when 1-2 LN metastases were identified, in contrast to only 22.4% in 2000 (41).

The prevalence of BCRL, that is the number of patients suffering from this disease, appears to be increasing. This is associated with a number of compounding factors, including increasing number of breast cancer diagnoses, higher patient survival rates, and greater global awareness of BCRL. In the United States alone, breast cancer incidence has increased 0.5% annually from 2010-2019 (42). A global forecast of breast cancer incidence, conducted by Xu *et al.*, has estimated a 32% increase in the age-stratified rate of breast cancer patients by 2050 in comparison to 2019, with a rate of 59.63 per 100,000 (43). By 2050, this will equate to an estimated 4.8 million cases worldwide, resulting in a continuing increase in the number of BCRL patients. This is of particular concern as long-term survival rates continue to improve; Cancer Research UK has reported that the 10-year net survival has increased 38% over the past 40 years (7). In the United States, breast cancer death rates have decreased 42% over the past 30 years (44).

With a predicted increase in BCRL patients over the next three decades, improvements in diagnostic methods are critical to improved patient care and management. A systematic review by O’Donnell *et al.* highlighted that limited guidelines exist to diagnose LE and lack high quality evidence from contemporary studies (45). Poor diagnostic standardisation impedes accurate evaluation and comparison of patient data and treatment outcomes. Improved incidence data will inform clinical practice, which in turn will facilitate earlier and more effective treatment for patients. Moreover, accurate incidence data will better inform healthcare policy on treatment plans that minimise adverse events.

Currently, a wide range of measurement methods are used to assess BCRL with considerable spread in the resulting incidence data (Fig. 6). The majority of assessment methods are circumference-based (Fig. 6b), largely driven by its clinical convenience (46). Our analysis suggests that circumference-based measurements lack diagnostic precision with large variances in incidence observed within each criterion’s patient population (Fig. 6d). This emphasises the poor correlation between criteria and incidence in circumference-based methods, owing in part to inconsistencies in the measurement techniques amongst healthcare systems. According to the International Society of Lymphology (ISL), however, volume-based measurements are the method of choice for patient assessment (47, 48). In contrast to circumference-based methods, volume measurements had improved reliability in this analysis, with statistically significant differences observed between criteria. The most commonly used criterion was an increase in volume of 10%, even though the ISL recommends that a minimum volume increase of 5% be utilised to identify LE since 2016 (49). A volume increase of 10% yielded the lowest BCRL incidence at 17.3%, and incidence increased by 50% to 25.5% when the criterion was reduced to 5%. This difference may directly impact whether a patient is diagnosed with subclinical lymphoedema, where early intervention is key to minimizing disease.

The ramifications of inconsistent recommendations for limb volume and circumference changes on incidence are clear. While indocyanine green lymphography, MRI, and lymphoscintigraphy are more sensitive for assessing early limb LE (50), their implementation in clinical practice is very limited, due to impracticality and expense. Therefore, the international lymphedema community must address the lack of standardisation in circumference and volume methods in order to improve patient management and outcomes. It is imperative that a consensus on the required number of serial measurements and their location, the methods to determine volume or circumference, and diagnostic criteria is reached.

In addition to a universally applied diagnostic regimen with good repeatability, improvements in patient data reporting are needed within cohort studies, with uniformly formatted databases of treatments used (and those not used) and full patient profiling. Inconsistencies in reporting practices of patient history and treatments received make estimating incidence a significant challenge. In 154 of the 184 qualifying studies in this analysis, it was not indicated what other treatments patients received beyond those that were the subject of each study. In the process of compiling and analysing the data for this meta-analysis, we do not assume a patient cohort received any treatments other than what is explicitly reported. It was for this reason, however, that treatments were grouped together by surgical or non-surgical methods. There was insufficient patient data to accurately investigate individual treatment protocols, particularly for non-surgical treatments (e.g. RT vs. HT). This led to a decrease in granularity in the ALND/SLNB groups that had both surgical and non-surgical treatments, as well as potential for bias within our results.

To avoid potential patient overlap when grouping by treatment type, it was necessary to exclude some incidence data from the analysis if patient cohorts from different groups were not mutually exclusive. The combination of incomplete patient data and excluded data may have an effect on the incidences reported, though the bias direction is unclear. It could contribute to some of the comparisons between treatment combinations failing to reach statistical significance, particularly in the ALND/SLNB groups that had both surgical and non-surgical treatments.

Further to this, our study did not consider studies that include preventive measures for BCRL, such as axillary reverse lymphatic mapping (ARM), lymphatic microsurgical preventive healing approaches (LYMPHA), or postoperative compression. This allowed for better assessment of the effect of treatment combinations in isolation on BCRL incidence. Furthermore, some of these preventive measures are not standard of care, such as ARM or LYMPHA, which limits their applicability in this analysis. A separate meta-analysis would be required to assess the effect of these interventions on BCRL incidence.

Our analysis provides data on the importance of axillary interventions (SLNB vs. ALND, number of LNs removed, and RT location) and patient BMI on BCRL incidence, but it also highlights the need for a paradigm shift in the approach to diagnosing and reporting BCRL. Widespread inconsistencies in diagnostic criteria and incomplete reporting practices lead to uncertainties in estimating any representation of incidence. Furthermore, given the bimodal effect of SLNB and ALND treatment on incidence, an overall incidence metric for BCRL, which is commonly utilised in patient and policy discourse, does not accurately reflect likelihood for the disease. These limitations greatly impede refined discourse of patient impact and, in turn, relevant policy development on breast cancer treatment and BCRL. In the short term, reporting SLNB and ALND-stratified incidence rates can provide a more robust basis for further investigations of incidence, shaping healthcare practices and policies, including doctor-patient discussions of relative procedural incidence escalation. Longer term, the international lymphoedema community must address these inconsistencies and shortcomings to improve patient care and outcomes.

### Future perspectives

Incidence only represents part of patient risk for BCRL, and overall risk should also be viewed in the context of the severity of the disease. By many measures, BCRL is grave, life-changing, and expensive. It has been recently shown that the severity of symptoms for BCRL, including pain and range of motion, depend on whether a patient received ALND or SLNB, with greater severity of symptoms for those who received ALND (51). In this context, it may be more appropriate to apply a risk matrix approach, in which risk is defined as the product of likelihood and severity scores. Given the bimodal nature of both BCRL incidence and severity, this embodies a more relevant representation of the relative risk of developing BCRL and will support the continued developments of means for preventing and curing BCRL.

## Data Availability

All data produced in the present study are available upon reasonable request to the authors

## References

1. Fu MR, Axelrod D, Haber J. Breast-cancer-related lymphedema: information, symptoms, and risk-reduction behaviors. J Nurs Scholarsh. 2008;40(4):341–8.

2. Jørgensen MG, Toyserkani NM, Hansen FG, Bygum A, Sørensen JA. The impact of lymphedema on health-related quality of life up to 10 years after breast cancer treatment. npj Breast Cancer. 2021;7(1):70.

3. Fu MR, Ridner SH, Hu SH, Stewart BR, Cormier JN, Armer JM. Psychosocial impact of lymphedema: a systematic review of literature from 2004 to 2011. Psycho-Oncol. 2013;22(7):1466– 84.

4. Sung H, Ferlay J, Siegel RL, Laversanne M, Soerjomataram I, Jemal A, et al. Global cancer statistics 2020: GLOBOCAN estimates of incidence and mortality worldwide for 36 cancers in 185 countries. CA Cancer J Clin. 2021;71(3):209–49.

5. Torgbenu E, Luckett T, Buhagiar MA, Chang S, Phillips JL. Prevalence and incidence of cancer related lymphedema in low and middle-income countries: a systematic review and meta-analysis. BMC Cancer. 2020;20(1):604.

6. Nardin S, Mora E, Varughese FM, D’Avanzo F, Vachanaram AR, Rossi V, et al. Breast cancer survivorship, quality of life, and late toxicities. Front Oncol. 2020;10:864.

7. Breast cancer survival statistics: Cancer Research UK; [Available from: https://www.cancerresearchuk.org/health-professional/cancer-statistics/statistics-by-cancer-type/breast-cancer/survival.

8. Lin Q, Yang T, Yongmei J, Die YM. Prediction models for breast cancer-related lymphedema: a systematic review and critical appraisal. Syst Rev. 2022;11(1):217.

9. McLaughlin SA, Brunelle CL, Taghian A. Breast cancer–related lymphedema: Risk factors, screening, management, and the impact of locoregional treatment. J Clin Oncol. 2020;38(20):2341–50.

10. McDuff SGR, Mina AI, Brunelle CL, Salama L, Warren LEG, Abouegylah M, et al. Timing of lymphedema after treatment for breast cancer: When are patients most at risk? Int J Radiat Oncol Biol Phys. 2019;103(1):62–70.

11. Iyigun ZE, Duymaz T, Ilgun AS, Alco G, Ordu C, Sarsenov D, et al. Preoperative lymphedema-related risk factors in early-stage breast cancer. Lymphat Res Biol. 2018;16(1):28–35.

12. Rupp J, Hadamitzky C, Henkenberens C, Christiansen H, Steinmann D, Bruns F. Frequency and risk factors for arm lymphedema after multimodal breast-conserving treatment of nodal positive breast cancer - a long-term observation. Radiat Oncol. 2019;14(1):39.

13. Gillespie TC, Sayegh HE, Brunelle CL, Daniell KM, Taghian AG. Breast cancer-related lymphedema: risk factors, precautionary measures, and treatments. Gland Surg. 2018;7(4):379–403.

14. Anita M, Andrea VB. Updates in surgical management of the axilla. Oncol. 2025;39(2):70–5.

15. Tinterri C, Gentile D, Gatzemeier W, Sagona A, Barbieri E, Testori A, et al. Preservation of axillary lymph nodes compared with complete dissection in T1-2 breast cancer patients presenting one or two metastatic sentinel lymph nodes: The SINODAR-ONE multicenter randomized clinical trial. Ann Surg Oncol. 2022;29(9):5732–44.

16. He L, Qu H, Wu Q, Song Y. Lymphedema in survivors of breast cancer. Oncol Lett. 2020;19(3):2085–96.

17. Asdourian MS, Swaroop MN, Sayegh HE, Brunelle CL, Mina AI, Zheng H, et al. Association between precautionary behaviors and breast cancer-related lymphedema in patients undergoing bilateral surgery. J Clin Oncol. 2017;35(35):3934–41.

18. Kapellas N, Demiri E, Lampropoulos A, Dionyssiou D. Genetic predisposition in cancer-related lymphedema: A systematic review. Lymphat Res Biol. 2022;20(5):478–87.

19. Newman B, Lose F, Kedda M-A, Francois M, Ferguson K, Janda M, et al. Possible genetic predisposition to lymphedema after breast cancer. Lymphat Res Biol 2012;10(1):2–13.

20. Kwan ML, Darbinian J, Schmitz KH, Citron R, Partee P, Kutner SE, et al. Risk factors for lymphedema in a prospective breast cancer survivorship study: The pathways study. Arch Surg. 2010;145(11):1055–63.

21. Anderson EA, Armer JM. Factors impacting management of breast cancer-related lymphedema (BCRL) in Hispanic/Latina breast cancer survivors: A literature review. Hisp Health Care Int. 2021;19(3):190–202.

22. Leray H, Malloizel-Delaunay J, Lusque A, Chantalat E, Bouglon L, Chollet C, et al. Body mass index as a major risk factor for severe breast cancer-related lymphedema. Lymphat Res Biol 2020;18(6):510–6.

23. Montagna G, Zhang J, Sevilimedu V, Charyn J, Abbate K, Gomez EA, et al. Risk factors and racial and ethnic disparities in patients with breast cancer-related lymphedema. JAMA Oncol. 2022;8(8):1195–200.

24. Fu MR. Breast cancer-related lymphedema: Symptoms, diagnosis, risk reduction, and management. World J Clin Oncol. 2014;5(3):241–7.

25. McLaughlin SA, Staley AC, Vicini F, Thiruchelvam P, Hutchison NA, Mendez J, et al. Considerations for clinicians in the diagnosis, prevention, and treatment of breast cancer-related lymphedema: Recommendations from a multidisciplinary expert ASBrS panel: Part 1: Definitions, assessments, education, and future directions. Ann Surg Oncol. 2017;24(10):2818–26.

26. Bucci LK, Brunelle CL, Bernstein MC, Shui AM, Gillespie TC, Roberts SA, et al. Subclinical lymphedema after treatment for breast cancer: Risk of progression and considerations for early intervention. Ann Surg Oncol. 2021;28(13):8624–33.

27. Rafn BS, Christensen J, Larsen A, Bloomquist K. Prospective surveillance for breast cancer– related arm lymphedema: A systematic review and meta-analysis. J Clin Oncol. 2022;40(9):1009–26.

28. DiSipio T, Rye S, Newman B, Hayes S. Incidence of unilateral arm lymphoedema after breast cancer: a systematic review and meta-analysis. Lancet Oncol. 2013;14(6):500–15.

29. Siotos C, Sebai ME, Wan EL, Bello RJ, Habibi M, Cooney DS, et al. Breast reconstruction and risk of arm lymphedema development: A meta-analysis. J Plast Reconstr Aesthet Surg. 2018;71(6):807–18.

30. Schünemann H, Higgins J, Vist G, Glasziou P, Akl E, Skoetz N, et al. Chapter 14: Completing ‘Summary of findings’ tables and grading the certainty of the evidence. In: Higgins J, Thomas J, Chandler J, Cumpston M, Li T, Page M, et al., editors. Cochrane Handbook for Systematic Reviews of Interventions 2025.

31. Norman SA, Localio AR, Potashnik SL, Simoes Torpey HA, Kallan MJ, Weber AL, et al. Lymphedema in breast cancer survivors: incidence, degree, time course, treatment, and symptoms. J Clin Oncol. 2009;27(3):390–7.

32. Rosenberger LH, Thomas SM, Plichta JK, Fayanju OM, Hyslop T, Greenup RA, et al. Decreasing rates of axillary lymph node dissections over time: Implications for surgical resident exposure and operative skills development. Am J Surg. 2019;218(4):786–91.

33. Cardoso F, Kyriakides S, Ohno S, Penault-Llorca F, Poortmans P, Rubio IT, et al. Early breast cancer: ESMO Clinical Practice Guidelines for diagnosis, treatment and follow-up. Ann Oncol. 2019;30(8):1194–220.

34. Ben-Dror J, Shalamov M, Sonnenblick A. The history of early breast cancer treatment. Genes. 2022;13(6).

35. Giuliano AE, Kirgan DM, Guenther JM, Morton DL. Lymphatic Mapping and Sentinel Lymphadenectomy for Breast Cancer. Ann Surg. 1994;220(3).

36. Giuliano AE, Ballman KV, McCall L, Beitsch PD, Brennan MB, Kelemen PR, et al. Effect of axillary dissection vs no axillary dissection on 10-year overall survival among women with invasive breast cancer and sentinel node metastasis: The ACOSOG Z0011 (Alliance) randomized clinical trial. JAMA. 2017;318(10):918–26.

37. Brackstone M, Baldassarre FG, Perera FE, Cil T, Chavez Mac Gregor M, Dayes IS, et al. Management of the axilla in early-stage breast cancer: Ontario Health (Cancer Care Ontario) and ASCO guideline. J Clin Oncol. 2021;39(27):3056–82.

38. Early and Locally Advanced Breast Cancer: Diagnosis and Treatment (NG101). National Institute for Health and Clinical Excellence Guidelines. Cardiff, UK: National Collaborating Centre for Cancer (UK); 2018.

39. Agrawal SK, Sharma H, Priya N, Saji AP, Phom HD, Sharma A, et al. Diagnostic performance and survival outcome following sentinel lymph node biopsy in breast cancer patients from a tertiary cancer centre in India. Ecancermedicalscience. 2022;16:1398.

40. Zhang J, Wang T, Yan C, Huang M, Fan Z, Ling R. Clinical practice satus of sentinel lymph node biopsy for early-stage breast cancer patients in China: A multicenter study. Clin Epidemiol. 2020;12:917–24.

41. Gou Z, Lu X, He M, Yu L. Trends in axillary surgery and clinical outcomes among breast cancer patients with sentinel node metastasis. The Breast. 2022;63:9–15.

42. Giaquinto AN, Sung H, Miller KD, Kramer JL, Newman LA, Minihan A, et al. Breast cancer statistics. CA Cancer J Clin. 2022;72(6):524–41.

43. Xu Y, Gong M, Wang Y, Yang Y, Liu S, Zeng Q. Global trends and forecasts of breast cancer incidence and deaths. Sci Data. 2023;10(1):334.

44. SEER Cancer Statistics Review, 1975-2018 [Internet]. National Cancer Institute. 2021. Available from: https://seer.cancer.gov/csr/1975_2018/.

45. O’Donnell TF, Jr., Allison GM, Iafrati MD. A systematic review of guidelines for lymphedema and the need for contemporary intersocietal guidelines for the management of lymphedema. J Vasc Surg Venous Lymphat Disord. 2020;8(4):676–84.

46. Cheville AL, McGarvey CL, Petrek JA, Russo SA, Thiadens SRJ, Taylor ME. The grading of lymphedema in oncology clinical trials. Semin Radiat Oncol. 2003;13(3):214–25.

47. The diagnosis and treatment of peripheral lymphedema: Consensus document of the International Society of Lymphology. Lymphology. 2003;36(2):84–91.

48. The diagnosis and treatment of peripheral lymphedema: 2020 Consensus Document of the International Society of Lymphology. Lymphology. 2020;53(1):3–19.

49. The Diagnosis and Treatment of Peripheral Lymphedema: 2016 Consensus Document of the International Society of Lymphology. Lymphology. 2016;49(4):170–84.

50. Mihara M, Hara H, Araki J, Kikuchi K, Narushima M, Yamamoto T, et al. Indocyanine green (ICG) lymphography is superior to lymphoscintigraphy for diagnostic imaging of early lymphedema of the upper limbs. PLoS One. 2012;7(6):e38182.

51. Che Bakri NA, Kwasnicki RM, Khan N, Ghandour O, Lee A, Grant Y, et al. Impact of axillary lymph node dissection and sentinel lymph node biopsy on upper limb morbidity in breast cancer patients: A systematic review and meta-analysis. Ann Surg. 2023;277(4):572–80.

